# Individual and community level factors associated with home delivery after adequate antenatal care visits: a multilevel analysis of the Ethiopian Demographic and Health Survey 2019

**DOI:** 10.1101/2023.09.21.23295888

**Authors:** Degefa Gomora, Girma Beressa, Kenbon Seyoum, Yohannes Tekalegn, Biniyam Sahiledengle, Daniel Atlaw, Neway Ejigu, Chala Kene, Telila Mesfin, Lillian Mwanri

**Author notes:** Corresponding Author [Degefa Gomora, ].

## Abstract

**Background:** Despite the reported increased antenatal care (ANC) utilization in Ethiopia, large numbers of women give birth at home without skilled personnel attendance, even after attending an adequate antenatal care schedule (≥ four antenatal visits) as recommended by the World Health Organization (WHO). This study aimed to assess individual and community-level factors associated with home delivery after adequate antenatal care visits in Ethiopia.

**Methods:** We analyzed the 2019 Ethiopian mini demographic and health survey data. A total weighted sample of 1,643 women who had full antenatal care visits for their last childbirth/index birth was included in the analysis. Model comparison was done by using intra-cluster correlation, median odds ratio, and proportional change in variance. A multivariable multilevel logistic regression analysis was conducted to identify the effect of individual and community-level factors on the outcome variable (home delivery). Adjusted odds ratios (AOR), along with a 95% confidence interval (CI) were used to estimate the strength of the associations.

**Results:** The intra cluster correlation (ICC) in the null model was 59%, showing that there was a significant difference in the prevalence of home delivery after adequate antenatal care at the community level, and the variability declined to 36.5% in the final model. Therefore, multilevel logistic regression model was utilized. At individual-level, secondary educational level was negatively associated with home delivery [AOR = 0.37; 95%CI: (0.17, 0.80)], and having a household of ≥ 5 members [AOR = 1.70; 95%CI: (1.09, 2.66)], poorest (vs. richest) wealth index [AOR = 6.98; 95%CI (2.89, 16.83)], poorer (vs. richest) wealth index [AOR = 2.77, 95%CI :(1.19,6.45)], and 2-3 birth order [AOR = 2.48; 95% CI(1.45, 4.21)] were associated with home delivery after attending the required adequate ANC visits. Community-level variables associated with home delivery after full antenatal care visits included: poor communities [AOR = 2.13; 95%CI (1.03, 4.40)], and living in a rural area [AOR = 2.74; 95%CI (1.19, 6.30)].

**Conclusion:** The findings of the current study indicate that in women who had the required number of ANC visits, having a larger household and poorest and poorer (vs. rich) wealth index, being the 2^nd^ or 3^rd^ birth, residing in a rural area, and living in poor communities were predictors of home delivery. Having a secondary education was supportive, against delivering at home. Strategies to improve household’s socioeconomic empowerment were recommended.

## Introduction

Globally, the majority of maternal deaths occur due to both pregnancy and childbirth, most commonly from complications of preventable causes such as infections and other circumstances and conditions that are preventable. In 2017, for example, over 800 women died per day of pregnancy and childbirth complications, of which 94% occurred in low and middle-income countries (LMICs) (1). According to the World Health Organization (WHO), the antenatal period is defined as the time between conception and birth. It is recommended that pregnant women in low and middle-income countries alone commence antenatal care (ANC) visits within the first four months after conception (2).

In Ethiopia, the rate of institutional delivery is low and complications related to pregnancy and childbirth are one of the leading causes of morbidity and mortality for women of childbearing age (3). In 2019 in Ethiopia, the maternal mortality rate was reported to be 412 per 100,000 live births, and the overall neonatal, infant, and under-five child mortality rate was 30, 43, and 55 deaths per 1000 live births respectively. Decreasing the proportion of home deliveries is a critical strategic effort to reduce maternal mortality rates. However, in 2019, more than half of the deliveries occurred at home in Ethiopia, where the majority of the women in the entire population lived in rural areas with low accessibility to health facilities. Furthermore, of the total home births in 2019 in Ethiopia, 60% of births were in a rural area, which is where the majority of neonatal and maternal deaths occurred. In Ethiopia, there is no infrastructure or health system to promote and support skilled birth attendance at home (4, 5).

Even though WHO released a 2016 statement recommending a minimum of eight ANC visits for pregnant mothers, Ethiopia applies the previous recommendation by WHO in 2002, indicating a minimum of four antenatal care visits for pregnant mothers (4, 6, 7). Despite an increase in the proportion of women receiving an adequate ANC follow-up (recommended four or more ANC visits) from 12% in 2005 to 43% in 2019, more than half of births to women who attended four or more ANC visits were delivered at home in 2019 (3, 4). In Ethiopia, per the prior WHO recommendation, four ANC visits are considered to be adequate ANC (7).

The proportion of home delivery in Ethiopia varies from 3.3% to 89.6% across the regions (8). Studies have revealed that home delivery was 48.53% in Ethiopia, and home deliveries were determined by factors including rural residence, absence of ANC, educational status, women’s age, and other socio-demographic variables (9,10).

Moreover, other studies in Ethiopia have indicated factors such as poor literacy, multi-gravida, husband’s educational status, not attending ANC, poor knowledge of obstetric complications, and walking time of greater than two hours to the health center as factors associated with home deliveries (11-13). Besides, previous primary studies indicated factors like not having exposure to media as one factor associated with home delivery (14-16).

Home delivery in Ethiopia was influenced by determinants including the quality of ANC, quality of information on birth preparation plans, not having health insurance, and individuals’ religion of residence. However, good wealth index and educational status have been reported to have a negative influence on home delivery (17, 18). Women’s beliefs were also a factor, especially when they considered home as a natural place for delivery and desired to deliver at home than at health facilities (13, 19).

In Ethiopia, skilled health professionals at health facilities give ANC. Although, it is widely acknowledged that ANC utilization can reduce home deliveries. However, the rate of home delivery is still the highest in low-income countries, including Ethiopia. Women who have had four or more antenatal visits at health facilities are expected to deliver children in the health facilities more often than women who had no antenatal care visits. However, more than half of births to women who attended four or more ANC visits were delivered at home in 2019 (3, 4). Most of the previous studies examining predictors of home delivery have not targeted women who had adequate (≥ four) ANC visits and/or did not take into account the clustering effects of the individual and community-level factors on home delivery. Thus, we aimed to assess individual and community-level factors associated with home delivery after adequate ANC visits in Ethiopia.

## Methods and Materials

### Study setting and Data source

The data source for this study was the 2019 Ethiopian Mini Demographic and Health Survey (EMDHS), the most recent and publicly available dataset. EMDHS is a nationally representative, the community-based cross-sectional household survey that was collected using multi-stage stratified cluster-sampling techniques. This study utilized the Kids Record (KR) file of the 2019 EMDHS data. Data (EMDHS) collection was carried out from March 21 to June 28, 2019 (4). The details of the sampling and data collection procedure are available on the DHS website (https://www.dhsprogram.com/).

### Sampling procedures

The 2019 EMDHS used a nationally representative sample that provided estimates at the national and regional levels as well as for urban and rural areas. The survey interviewed 8,855 women of reproductive age (15-49 years) from a nationally representative sample of 8,663 households. In this study, women were included if they had one or more births in the five years preceding the survey, had adequate (at least four) ANC visits, and had a record of the place of the more recent birth. Women who had ≤ 3 ANC visits, women who had another place of delivery (rather than the respondent’s home, government or private health institution, their own home), or women who were visitors (not live in the respondents’ home permanently) were excluded. From a total weighted sample of 3,927 women who had last live births in five years (coded in the survey data as “midx = 1”), a weighted sample of 1,643 women who had at least four ANC follow-ups were included in the analysis (Figure. 1).

**Figure 1.**
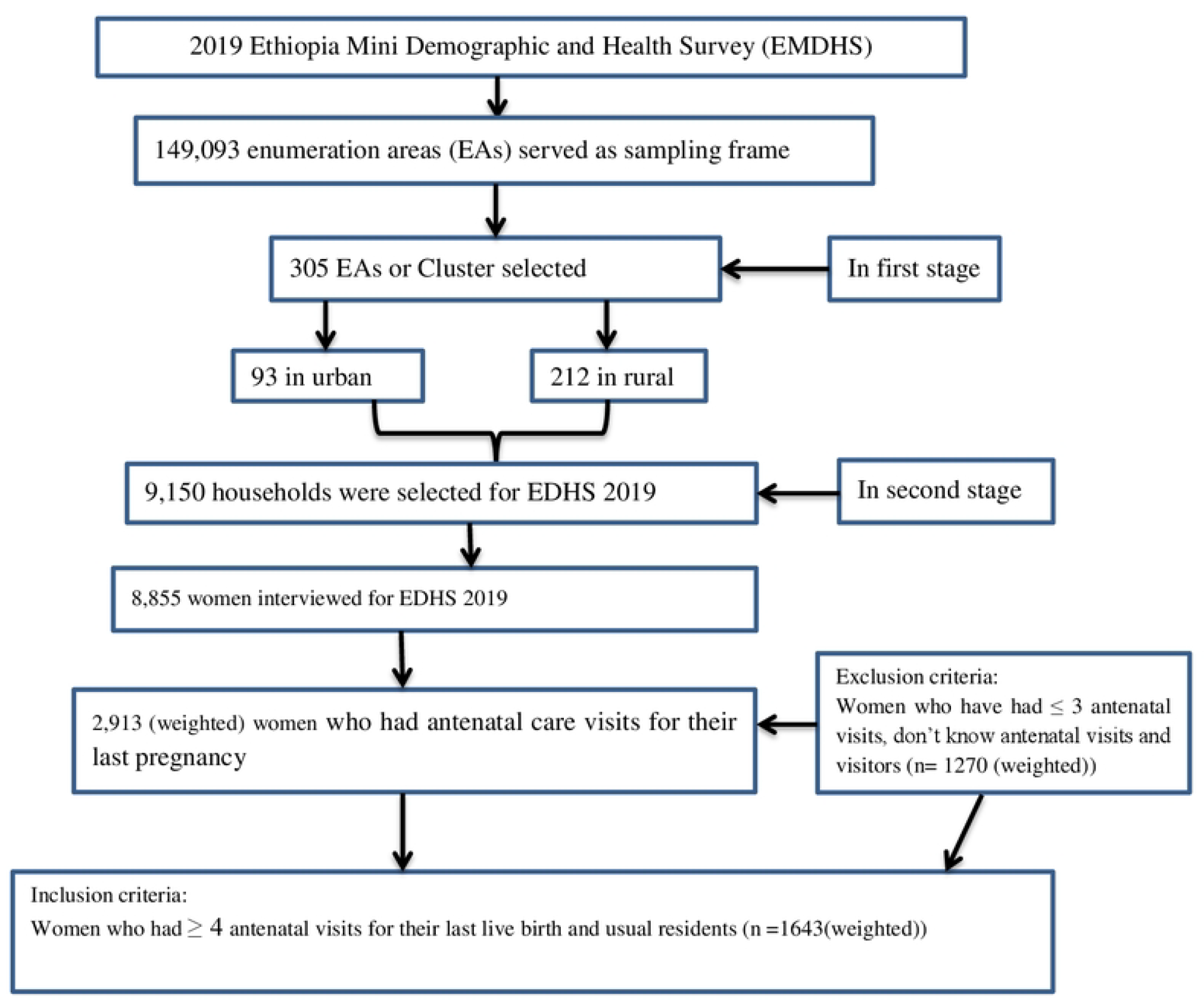
Sampling procedures for individual and community level determinant of delivery after adequate antenatal care visits, EMDHS 2019.

### Outcome variables

The place of delivery among mothers who had adequate ANC visits (at least four ANC visits at a health facility) was as follows: Home delivery was assigned 1, and health institution delivery was assigned 0. Home delivery is a delivery that was not attended by doctor, nurse, midwife, health officer, and health extension worker (4).

### Explanatory variables

Individual level and community-level factors included in this study were as follows: (i) Individual-level factors include; women’s highest educational level, women’s age, marital status of the women, religion, number ≤5 years old children in household, sex of household head (is house is led by the male or female?), wealth index (household level wealth index calculated from wealth index indices and categorized into poorest, poorer, middle, richer, and richest), sons or daughters who have died (sons or daughters born alive but later died), age of women at first birth, birth order of the most recent birth; the number of household members (includes all person that lives in the household, but not visitors), number of living children, the timing of first antenatal check (month of pregnancy in which first prenatal check was received).

(ii)Community-level factors were of two types: integral variables (community type) and derived (aggregates) variables. Community level variable was generated by aggregating the individual characteristics with which interest in a cluster. The aggregate was computed using the proportion of a given variable’s subcategory; we were concerned about in a given cluster. Since the aggregated value for generated variables was not normally distributed. It was categorized into groups based on the median values (20). Community-type community-level factors included in this study were the place of residence “urban or rural, and the regions were also one of the community variables further categorized as agrarian (Tigray, Amhara, Oromia, Southern Nations, Nationalities and People’s region (SNNPR)), pastoralists (Afar, Benshangul-Gumuz, Gambela, and Somali), and metropolises (city administrations) (Harari, Addis Ababa, and Dire Dawa).

The aggregated type of the community-level variables included in this study was the community wealth index, which indicates the proportion of women in the two lower levels (poorest and poorer). Wealth index components in the community and categorized using the median split (median=14.3%) as high community wealth and low community wealth for the proportion above and below the median, respectively, and the community education which was defined as the proportion of women who attended primary and secondary education or above within the cluster. This proportion was divided into two using the median value (median=63.6%), categorized as low for the proportions, below the median value, and high for the proportions above the median value within the cluster (Table 1).

**Table 1 Individual and community variables used in the analysis**

### Statistical analysis

Due to the dichotomous nature of the place of delivery, a two-level mixed effect logistic regression analysis was conducted. Individual and cluster (community) levels made up the two levels. The log of the likelihood of giving birth at home was thus represented using a two-level multilevel model as follows. 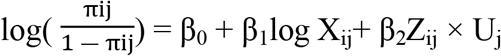 Where, i and j are the level 1 (individual) and level 2 (community) units, respectively; X and Z refer to individual and community-level Variables, respectively, πij is the probability of home delivery for the i^th^ woman in the j^th^ community; the β′s are the fixed coefficients. Therefore, for every one unit increase in X or Z (a set of predictor variables), there is a corresponding effect on the probability of the woman having a home delivery. Whereas, β0 is the intercept the effect on the probability of a woman to have a home delivery in the absence of influence from predictors; and Uj shows the random effect (effect of the community on whether a woman has to have a home delivery for the j^th^ community. Four models were fitted to estimate both the fixed effects of the individual and community-level factors and the random effects of between-cluster variation.

Data analysis was done by STATA 17, and sample weights were applied to adjust for non-proportional allocation of samples and possible differences in response rates across regions included in the survey.. No missing data was present. Due to the hierarchical nature of the EMDHS data and the presence of intra-class correlation (ICC), multilevel logistic regression analysis was used. Bivariate and multivariate multilevel logistic regression analyses were conducted to determine the independent effect of individual and community level variables on the dependent variable. Independent variables with a p-value of less than 0.25 during bivariate multilevel logistic regression analyses were considered for multivariable multilevel logistic regression analysis. The results of the fixed effects model were presented as OR along with 95% confidence intervals (CIs).

An adjusted odd ratio (AOR) along with 95% CIs was computed to estimate the strength of the associations between factors associated with place of delivery among women who had full ANC visits. A p-value less than 0.05 determined to be statistically significant. ICC, proportional change in variance (PCV), and median odd ratio (MOR) measured the random effects (variation of effects), which measure the variability between clusters in the multilevel models (21-23). ICC explains the cluster variability, while MOR can quantify unexplained cluster variability (heterogeneity). MOR converts cluster variance into an OR scale. In the multilevel model, PCV can measure the total variation due to factors at the community and individual level (22).

The ICC, PCV and MOR were determined using the estimated variance of clusters using the following formula. 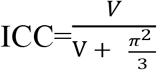 Where, V denotes community variance; and 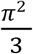 denotes individual level variance that is fixed for log distribution (equal to 3.29). 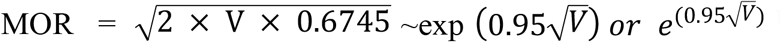; Where V is the estimated variance of clusters and 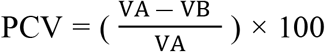 Where VA = variance of the initial (null) model, VB = variance of the model with more terms ((21, 22). Model fits were assessed using log likelihood (LL), deviance, and Akaike information criterion (AIC). LL, AIC, and deviance was used to estimate the goodness of fit of the adjusted final model in comparison to the preceding models (individual and community level model adjustments). The LL, AIC, and deviance values for each subsequent model were compared and the model with the highest value of LL and lowest value of deviance and AIC was considered the best-fit model.

Model comparison was conducted for the null/model 1 (model without explanatory variables), model 2 (model adjusted for individual level factors), model 3 (model adjusted for community level factors), and model 4 (final model adjusted for both individual and community-level factors). A variance inflation factor (VIF) was conducted to check for the presence of multicollinearity among exposure variables.

### Operational definitions

#### Adequate antenatal care (ANC) visits

If women had ≥ 4 antenatal visits at health facilities (24-26)

### Sociodemographic characteristics of the respondents

This study analyzed the weighted sample of 1,643 women who had full ANC visits at health facility, and who had an alive birth in the five years preceding the EMDHS 2019. Thirty-nine percent (n=633) of the women had no formal education, while 14.5% (n=240) had secondary educational. The households of more than half 60.7% (n=998) women had ≥ 5 household members. Almost ten percent 9.8% (n=162) of the women were in the poorest wealth index while about 18.5% (n=304) were in the poorer wealth index. More than half 55.1% (n=904) of the women in the community had a higher proportion of primary and above educational level (Table 2).

**Table 2 Sociodemographic characteristics of women who had adequate ANC visits in Ethiopia, 2019 EMDHS, (N**^**a**^ **= 1643)**

### Rate of home delivery

Overall, the proportion of home deliveries among women who had adequate ANC visits (≥4 visits) was 25.6% (95% CI: 23.55, 27.78). More than one-fourth, 26.08% (n=410) of women had 4-7 antenatal visits, and nearly fifteen percent, 14.9% (n=10) of the women had greater than or equal to eight antenatal visits in 2019. The chi-square analysis of the place of delivery with the grouping of ANC visits showed that the proportion of home delivery was significantly reduced from 26.1% among women who had 4-7 antenatal visits to 14.9% among those who had ≥8 antenatal visits (*X*2 = 14.56,p < 0.001).

#### Bivariate analysis

Individual level variables including highest educational level, age at 1st birth, sex of household head, sons or daughters who have died, wealth index combined, birth order, number of household members, number of living children, and community level variable including community wealth index, residence community education, and region were significant at p-value ≤ 0.2 during bivariate analysis. However, the variables with > 0.2 p values in bivariate analysis (religion, age of respondents, current marital status, timing of first antenatal care visits) were excluded from the models. On the other hand, numbers of living children were excluded due to similar concepts and collinearity effects with birth order (Table 3).

**Table 3.** Bivariate multilevel logistic regression analysis for individual and community level factors associated with home delivery after adequate ANC visits, EMDHS 2019.

| Place of delivery |  |  |  |  |  |
| --- | --- | --- | --- | --- | --- |
|  |  | Health facility Count (%) | Home Count (%) | COR(95%CI) | P value |
| **Individual level factors** |  |  |  |  |  |
| Highest educational level | No education | 409(64.7) | 223(35.3) | Ref |  |
|  | Primary | 494(75.5) | 160(24.5) | 0.60(0.41,0.87) | 0.008 |
|  | Secondary | 214(89.3) | 26(10.8) | 0.22(0.10,0.48) | 0.001 |
|  | Higher | 105(90.1) | 12(9.9) | 0.20(0.05,0.78) | 0.021 |
| Age at 1<sup>st</sup> birth | < 18 | 401(65.8) | 208(34.2) | 2.01(1.40,2.88) | 0.001 |
|  | ≥ 18 | 822(79.4) | 213(20.6) | Ref |  |
| Sex of household head | Male | 1052(73.1) | 387(26.9) | 1.85(1.05,3.24) | 0.032 |
|  | Female | 171(83.4) | 34(16.6) | Ref |  |
| Sons or daughters who have died | No son and daughter died | 1033(76.6) | 316(23.4) | Ref |  |
|  | Son/s or daughter/s died | 167(63.6) | 96(36.4) | 1.87(1.20, 2.92) | 0.005 |
|  | Sons and daughters died | 22(71.5) | 9(28.5) | 1.30(0.46,3.69) | 0.618 |
| Number of household member | <5 | 556(86.2) | 89(13.8) |  |  |
|  | ≥5 | 666(66.8) | 332(33.3) |  |  |
| Wealth index | Poorest | 74(45.6) | 88(54.4) | 10.20(5.16,20.37) | 0.001 |
|  | Poorer | 196(64.4) | 108(35.6) | 4.75(2.46, 9.16) | 0.001 |
|  | Middle | 191(65.8) | 99(34.2) | 4.46(2.31, 8.62) | 0.001 |
|  | Richer | 256(79.4) | 67(20.7) | 2.23(1.12, 4.45) | 0.022 |
|  | Richest | 506(48.8) | 59(10.4) | Ref |  |
| Birth order | 1 Birth order | 364(89.7) | 42(10.3) | Ref |  |
|  | 2-3 Birth order | 455(75.9) | 144(24.1) | 2.75(1.46, 5.19) | 0.002 |
|  | 4+ Birth order) | 403(63.2) | 235(36.8) | 5.06(2.75, 9.29) | 0.001 |
| **Community level factors** |  |  |  |  |  |
| Community education | Low | 623(67.7) | 297(32.3) | 2.31(1.54, 3.46) | 0.001 |
|  | High | 600(82.9) | 124(17.1) | Ref |  |
| Community Wealth index | Low | 647(87.6) | 91(12.4) | 0.25(0.16,0.38) | 0.001 |
|  | High | 575(63.6) | 329(36.4) | Ref |  |
| Residence | Urban | 507(85.5) | 86(14.5) | Ref |  |
|  | Rural | 715(68.1) | 335(31.9) | 2.75(1.66, 4.57) | 0.001 |
| Region | Agrarian | 1056(72.7) | 397(27.3) | 10.61(5.85, 19.26) | 0.001 |
|  | Pastoralist | 51(72.2) | 20(27.8) | 10.89(5.58, 21.24) | 0.001 |
|  | Metropolis | 115(96.6) | 4(3.4) | Ref |  |

#### Model comparators

A multilevel logistic regression model was utilized in this study due to the intra-cluster correlation (ICC) in the null model being 59%, showing that there was a significant difference in the prevalence of home delivery after adequate ANC at the community level. Moreover, the variability declined to 37% in the final model (model adjusted for both individual and community-level factors). The null model showed that there was significant variability in the odds of having a home delivery after adequate ANC visits (variance = 4.73, p < 0.001). The median odds ratio (MOR) indicated that having a home delivery after adequate ANC was attributed to community-level factors. In the null model, MOR was 2.53, indicating that there was variation between clusters. Individuals in the cluster with the higher odds of having a home delivery after adequate ANC visits had 2.53 times the odds of giving birth at home after adequate ANC visits as individuals in the cluster with the lower odds of giving birth at home after adequate ANC visits. Finally, the final model with the highest log-likelihood (-619.11), lowest AIC (1280.23), and lowest deviance (1238.22) was selected (Table 4).

**Table 4.** Random effect parameters and model comparators for the study individual and community level factors associated with home delivery after adequate ANC visits, 2019 EMDHS.

| Comparator | Null/ model 1 | Model 2 | Model 3 | Model 4 |
| --- | --- | --- | --- | --- |
| Community variance(SE)<br>(95%CI) | 4.73(0.9295)<br>(3.21,6.95) | 1.68(0.393)<br>(1.06,2.66) | 2.32(0.504)<br>(1.52,3.55) | 1.89(0.442)<br>(1.20, 2.99) |
| ICC (%)<br>(95%CI) | 59.0<br>(49.42,67.87) | 33.8<br>(24.39,44.67) | 41.4<br>(31.57,51.93) | 36.5<br>(26.67, 47.61) |
| PCV (%) | Reference | 64.48 | 50.95 | 60.04 |
| MOR | 2.53 | 1.51 | 1.77 | 1.60 |
| Log-likelihood(LL) | -715.84 | -628.48 | -659.17 | -618.08 |
| AIC | 1435.682 | 1288.95 | 1332.36 | 1278.167 |
| BIC | 1446.456 | 1375.14 | 1370.069 | 1391.295 |
| Deviance | 1431.68 | 1256.96 | 1318.34 | 1236.16 |

**Notes:** **AIC:** Akaike information criteria; **ICC:** Intra cluster correlation; **MOR:** Median odds ratio; **PCV:** Proportional change in variance; **Null/model 1:** model without explanatory variables; **Model 2:** Model adjusted for individual level factors; **Model 3:** Model adjusted for community level factors; **Model 4:** Final model adjusted for both individual and community level factors

#### Determinants of home delivery after adequate antenatal care visits

The final multivariable multilevel model/model 4 (model adjusted for both individual and community factors) revealed household members, wealth index, and birth order. While community wealth index and residence were significantly associated with home delivery after adequate ANC visits, and having a higher education was significantly associated with home delivery. Among women who had at least four ANC visits (adequate ANC), those who had a secondary educational level were less likely to deliver at home as compared to those who had no formal education [AOR = 0.37; 95%CI: (0.17, 0.80)]. Besides, the odds of having a home delivery after having an adequate ANC visit were higher among women who had ≥ 5 household members compared to those who had < 5 households [AOR = 1.70; 95%CI: (1.09, 2.66)].

Additionally, the likelihood of having a home delivery after having adequate ANC visits was higher among women who were in the poorest wealth index as compared to those who were in the richest wealth index [AOR = 6.98; 95%CI (2.89, 16.83)]. The odds of having a home delivery after having adequate ANC visits were higher among women whose last birth was in a 2 to 3 birth order when compared to those whose last birth was in the first birth order [AOR = 2.48; 95% CI(1.45, 4.21)]. Among community-level variables, women from highly poor communities were more likely to deliver at home after having adequate ANC visits compared to women from low-poor communities [AOR = 2.13; 95%CI (1.03, 4.40)]. Women from rural residences were more likely to deliver at home after having adequate ANC visits when compared to urban women [AOR = 2.74; 95%CI (1.19, 6.30)] (Table 5).

**Table 5. Multivariable multilevel logistic regression analysis for individual and community level factors associated with home delivery after adequate ANC visits, EMDHS 2019.**

## Discussion

Antenatal care (ANC) attendance, especially in LMICs is one of one of the significant contributors to reducing infant and maternal morbidity and mortality, and has been acknowledged to be effective in monitoring and managing any complications during pregnancy and childbirth (27). The 2019 EMDHS was conducted in the last five years of 2019 and in 2002; the WHO recommendation is for all pregnant women to have at least four ANC visits, with additional appointments should complications be detected during the course of the pregnancy. A number of factors can influence pregnant women’s attendance to ANC services and a place where the woman would deliver their baby (28). This study examined the individual and community level factors associated with home delivery among women who had at least four ANC visits for their last birth preceding the Ethiopian 2019 mini demographic health survey. In Ethiopia, per the prior WHO recommendation, four ANC visits is considered to be adequate ANC (7).The analysis works among women who had at least 4 ANC and it might in support to recommend whether the new WHO recommendation is still important in Ethiopia. The analysis showed that the between-cluster variability declined over successive models, from 59% in the empty model to 36.5% in the final model that was adjusted for both individual and community level factors.

Thereafter, the final model revealed that different individual and community factors were responsible for home deliveries after having adequate ANC visits (at least four ANC visits). From individual-level factors, low educational level, the number of household members, wealth index, and birth order were significantly associated with home delivery after having adequate ANC visits in Ethiopia. On the other hand, community-level wealth index and place of residence were community factors associated with home delivery.

Therefore, after controlling for individual and community factors, among women who had adequate ANC visits the odds of having a home delivery were lower among women who had a secondary education level than among women who had no educational level. The findings of this study were consistent with those of studies conducted in Ethiopia (18, 29, 30) and study in India (31).

Educated women are more likely to use healthcare facilities in Ethiopia (32, 33). Overall, this study reaffirms the significance of educating women. Education has been used as a vehicle for national socioeconomic development (34), as well as for individual advancements including in decision-making. It is plausible to assume that educated women would have a better understanding of why they should need skilled care attendance during delivery. Additionally, educated women can travel outside the home, assuming that their education would have placed them in a better socioeconomic situation to afford transport and may have a greater decision-making capability and autonomy (35) in seeking health services (33, 36, 37). Yet, this study showed that the odds of having home delivery after a full antenatal care visit were higher among women who had ≥5 household members than those who had <5 household members. This is possibly due to the family responsibilities of having a large family that might have interfered with travel plans or a stay in a health facility. Additionally, the likelihood of having a home delivery after having full ANC visits was higher among women who were in the poorest wealth index as compared to those who were in the richest wealth index. This study’s findings are in line with findings from studies conducted in Ethiopia (29, 38), and supported by the studies carried out in Afghanistan, and Mozambique (39, 40). This might be because 91.7% (n=220) of the poorest women were from rural areas where there was low access to health facilities and low educational levels, due to poor access to education opportunities, especially in Ethiopia and further increases home deliveries (41, 42).

Moreover, the odds of having home delivery after having adequate antenatal care visits were higher among women whose last birth was in 2 to 3 birth orders when compared to those whose last birth was in the first birth order. This finding is consistent with findings from other parts of Ethiopia (19, 43). Similarly, this finding is supported by findings from Ghana and Pakistan (44-46). However, evidence exists that the high likelihood of home delivery among multiparous women could also be due to socioeconomic burden. (32, 45, 47, 48), where these women could, for example not be able to afford transport with large families needing more resources for living.

Among community-level variables, women from the high-poor community were more likely to deliver at home after having adequate ANC visits compared to the low-poor community. This finding is comparable with findings from Ethiopia (18, 49), Bangladesh (45), Nigeria (50), and other studies (Delta state Nigeria, Nigeria, and India) (51-53). This might be because home delivery may be the norm in communities with high concentrations of poverty, and the majority of poor communities were found in rural areas in Ethiopia where there is low accessibility to health facility might prompt women to deliver at home.

Like other studies (8, 32, 49, 54, 55), we found that women from rural areas were more likely to deliver at home when compared to urban women. The possible reason might be due to low access to health facilities as compared to urban residents (8), The other reason might also due to distance to the health facilities, ambulance delays due to distance and difficult roads, inaccessibility of information about the advantage of institutional delivery and low educational status of women from rural areas (56).

Overall, the findings showed that delivering at home after adequate ANC in Ethiopia was common with over a quarter of women (25.6%) reporting having home delivery. The determinants of home delivery were consistent with the WHO Social Determinants of Health Framework (57). The Social Determinants of Health Framework recognizes poor access to health services, low levels of education, and poverty as significant determinants of health.

### Strength and limitations of the study

This is the first study to assess the determinants of individual and community level determinants of home delivery among women who had adequate ANC (at least four ANC visits at a health facility) in Ethiopia using a nationally representative cross-sectional study from 2019 EMDHS (Mini Demographic and Health Survey) data. This is the first study that assessed both individual-level and community-level factors related to home delivery among women who have at least 4 ANC visits in Ethiopia. The study used a multilevel logistic regression model, which enhances the accuracy of estimates since EMDHS data has a hierarchical nature. Due to the secondary nature of the data, a limited number of variables were included, in this analysis. Due to the cross-sectional nature of the data, a cause-effect relationship between the outcome and independent variables could not be established.

## CONCULUSION

Home delivery is a significant issue not only in Ethiopia but also in other developing countries. Having a secondary education, a larger number of household members, the poorest, and poorer wealth index, a lower birth order, rural residence, and being from a highly poor community were predictors of home delivery after having adequate ANC visits (at least four ANC visits at a health facility). To improve maternal and child outcomes in Ethiopia, it is necessary to address broader socioeconomic determinants in the community including individual and community factors that were observed in the current study, acknowledging that home delivery can be unsafe, especially for pregnant women with complications (58). Notably, strategies to improve women’s levels of education and economic empowerment are indispensable. Increasing the health services for all women, especially for women living in rural areas where low accessibility to the health facility. Additionally, increasing the antenatal care visits (at least 8) is recommended.

## Acknowledgements

The authors acknowledge the international major DHS program for giving permission EMDHS 2019 for data.

## Ethics Statement

Data from the 2019 Ethiopian Demographic and Health Survey were used in this study as a secondary analysis. Following submission of a proposal to the DHS Program, ethical approval was obtained, and the International Review Board of the Demographic and Health Surveys (DHS) (Reference No. DHS ICF 13/2023), program data archivists confirmed that download of the dataset for this study was permitted. The secondary data utilized in this study were collected from publically available sources and did not contain any information that could be used to personally identify study participants. When collecting data for the MEDHS 2019, anonymity was maintained to protect data confidentiality. For this particular investigation, there was no formal ethical review necessary. Information gleaned from the data set was made available to anyone else.

## Data Availability

The dataset used for the preparation of this manuscript is available from http://www.dhsprogram.com/, and anyone can access it through an online request as an authorized user. The corresponding author (Degefa Gomora) prepared the data that was used for the preparation of this manuscript, which can be shared if required.

## Consent for publication

NOT APPLICABLE

## Authors’ contributions

DG conceptualizes the idea, study design, execution, and acquisition of data, analysis, interpretation of the result, drafted the manuscript, revise, or critically reviewed the article. KS contributed to obtaining data, statistical analysis, interpretation of the results, and revision of the manuscript. GB, KS, YT, BS, DA, NE, CK, TM, and LM contributed to the conceptualization of the study, analytical strategy, and interpretation of results reviewed the first draft, and drafted the subsequent versions of the manuscript. All authors read and approved the final manuscript. All authors have agreed on the journal to which the article be submitted. Agree to take responsibility and be accountable for the contents of the article.

## LIST OF ABREVIATION

ANC: Antenatal Care;
AOR: Adjusted Odds Ratio;
CI: Confidence Interval;
COR: Crude Odds Ratio;
EA: Enumeration Area;
EMDHS: Ethiopian Mini Demographic Health Survey;
ETB: Ethiopian Birr;
EHSTP: Ethiopian Health Sector Transformation Plan;
WHO: World Health Organization.

